# Analysis of the effect of proton pump inhibitors on the course of common COVID-19

**DOI:** 10.1101/2020.06.07.20124776

**Authors:** Xiao-Yu Zhang, Hai-Bing Wu, Yun Ling, Zhi-Ping Qian, Liang Chen

**Author notes:** **Correspondence:** Liang Chen, Xiao-Yu Zhang. **E-mail addresses** Xiao-Yu Zhang, Hai-Bing Wu, Yun Ling, Zhi-Ping Qian, Liang Chen.

## Abstract

**Background/aims:** To evaluate the effect of proton pump inhibitors on the course of common COVID-19.

**Methods:** Clinical data of common COVID-19 patients admitted to the Shanghai public health clinical center for treatment from January 20, 2020 to March 16, 2020 were collected. A retrospective study was conducted and the patients were divided into two groups according to whether they used proton pump inhibitors or not. The differences in SARS-CoV-2 clearance and hospital stay between the two groups were compared by univariate and multivariate analyses.

**Results:** A total of 154 COVID-19 common cases were included in this study, including 80 males (51.9%), 35 patients (22.7%) in the proton pump inhibitors group, and 119 patients (77.3%) in the control group. In the proton pump inhibitors group and the control group, the duration of SARS-CoV-2 clearance were 7(6-9) and 7(6-11) days, and the duration of hospital stay was 21(16-25) and 20(15-26) days, respectively. There was no significant difference between the two groups in the cumulative incidence of SARS-CoV-2 clearance and the cumulative incidence of discharge, and the same after Propensity Score Match, all P > 0.05. Multivariate analysis suggested that chronic gastropathy prolonged the duration of SARS-CoV-2 clearance, the HR was 20.924(3.547-123.447). Hypertension, chronic obstructive pulmonary disease, chronic liver disease and malignant tumor all increased the duration of hospital stay for COVID-19, and the HR were 1.820 (1.073-3.085), 4.370 (1.205-15.844), 9.011 (2.681-30.290) and 5.270 (1.237-22.456), respectively; the duration of hospital stay in COVID-19 patients was shortened by SARS-CoV-2 clearance, and the HR was 0.907 (0.869-0.947); all P < 0.05.

**Conclusion:** Proton pump inhibitors use have no effect on the prolonging or shortening of the course of adults hospitalized with COVID-19.

## Introduction

The disease burdens of coronavirus disease 2019 (COVID-19), caused by severe acute respiratory syndrome coronavirus-2 (SARS-CoV-2), have been continuously increasing. [1]As of 6:50 p.m. Edt on 22 May 2020, 4,995,996 confirmed cases of COVID-19, including 327,821 deaths, have been reported to WHO.[2]Some COVID-19 patients have obvious gastrointestinal symptoms [3-4], and some basic diseases and therapeutic drugs may cause damage to the gastric mucosa; which can lead to PPIs being used to varying degrees in the treatment of COVID-19.

However, proton pump inhibitors (PPIs) have been found in several clinical studies to increase the incidence of community-acquired pneumonia or hospital-acquired pneumonia.[5-12] On the other hand, studies have showed that PPIs did not increase the incidence of pneumonia. They believed that the increased incidence of pneumonia caused by PPIs was due to confounding factors, and PPIs itself was not an independents factor affecting the occurrence of pneumonia.[13-16]

Basic studies have shown that PPIs can effectively inhibit virus-specific serine proteases and thus play an antiviral role.[17] And H+/K+-ATPase inhibitors can be tried to treat viral infection and respiratory disease.[18] However, other findings have been reported that gastrointestinal microorganisms modulate immune responses at distant mucosal sites and have the ability to significantly influence mortality during respiratory viral infection.[19]

It is not clear whether PPIs will have a significant impact on COVID-19. The purpose of this study was to analyze whether PPIs has relation with SARS-CoV-2 clearance and length of hospital stay in COVID-19.

## Methods

### Study design

A retrospective cohort study design was conducted in Shanghai Public Health Clinical Center affiliated to Fudan University, a designated tertiary teaching hospital for the treatment of SARS-CoV-2 infection. Patients with common COVID-19 confirmed diagnosis during the first three days in the hospital from January 20, 2020 to March 16, 2020 were included, and those cases would be followed up to discharge.

Informed consents of patients were obtained for diagnosis and treatment, and the study protocol was approved by the Shanghai Public Health Clinical Center Clinical Committee. All the data received Institutional Review Board (IRB) approval by the Ethics Committee. The IRB number was YJ-2020-S015-01.

### Patients and definitions

A total of 298 adults admitted were diagnosed with common COVID-19 in the first three days after admission.

common COVID-19 met following criteria: (1) the patients have epidemiological contact history or clinical manifestations; (2) pneumonia can be seen on imaging; (3) the SARS-CoV-2 nucleic acid test was positive by real-time fluorescence RT-PCR; (4) respiratory rate > 12 times/min and < 30 times/min, oxygenation index > 300mmHg, oxygen concentration > 93% in resting state; (5) no shock and organ failure.

SARS-CoV-2 clearance met following criteria: the SARS-CoV-2 nucleic acid test was negative for two consecutive pharyngeal swabs, sampling time interval at least 24 hours.

Discharge met following criteria: (1) body temperature returned to normal for more than 3 days; (2) respiratory symptoms improved significantly; (3) pulmonary CT scan showed basic absorption of acute exudative lesions; (4) the patients obtained SARS-CoV-2 clearance.

### Inclusion and exclusion criteria

A total of 154 cases were enrolled in the analysis. Of the 144 cases excluded, 27 cases were given PPIs for less than 3 days before the SARS-CoV-2 clearance, 12 cases had no nucleic acid test within the first three days after admission, 104 cases lacked continuous detection data required for the study design, and 1 case died without exposure to PPIs. See figure 1 for details.

**FIG 1.**
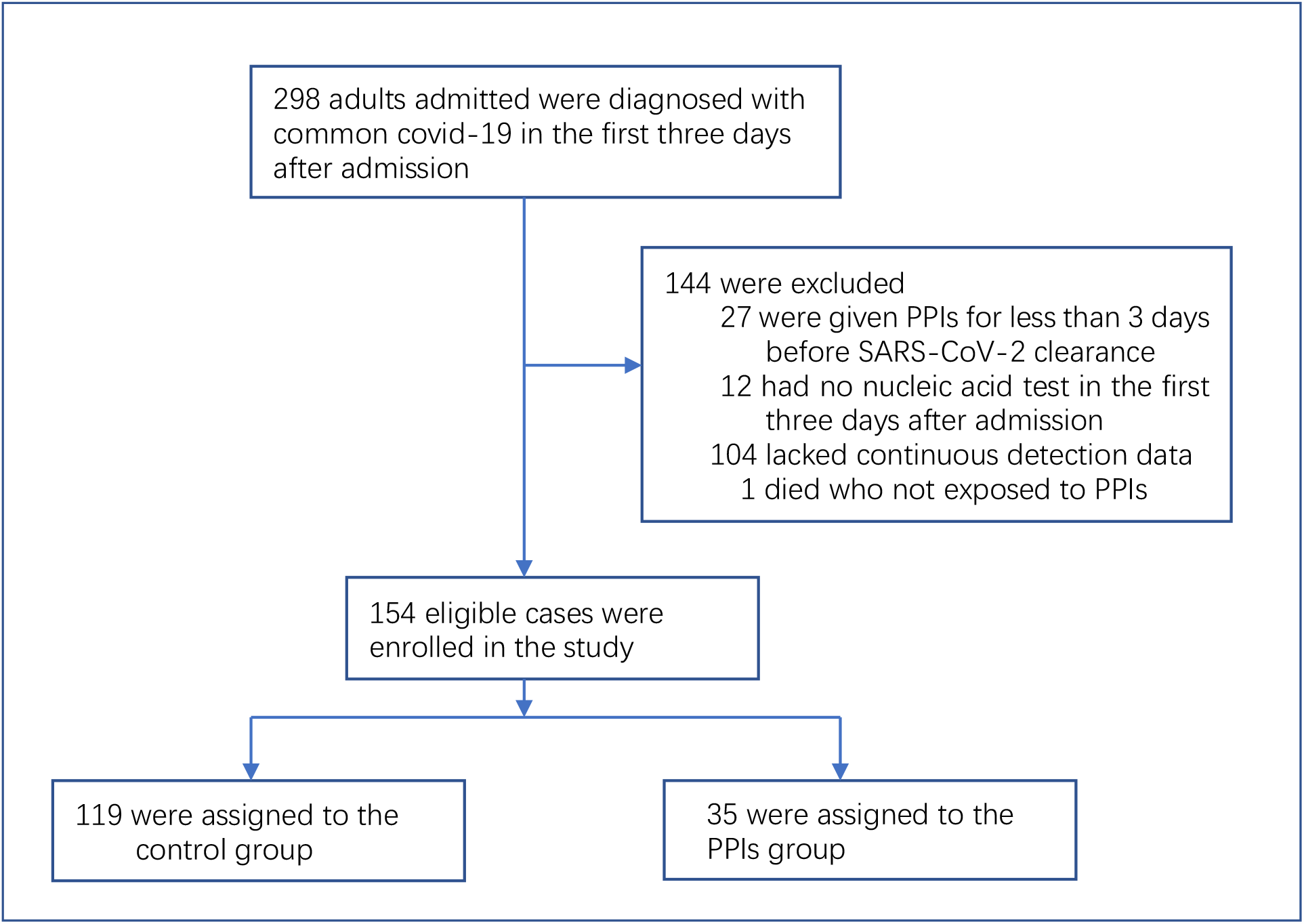
Study Cohort.

Case inclusion criteria: (1) cases were diagnosed with common COVID-19 within first three days after admission; (2) eligible cases were man and non-pregnant women; (3) age > 18 years old; (4) the SARS-CoV-2 nucleic acid were tested positive for the cases during the first three days after hospitalization; (5) the nucleic acid test of SARS-CoV-2 was continuous; (6) patients in the PPIs group were those who had used PPIs for at least 3 days before the SARS-CoV-2 clearance, while patients in the control group were those who had not used drugs to inhibit gastric acid secretion; (7) the daily dose of omeprazole was 20mg or 40mg, or the daily dose of rabeprazole was 20mg.

Case exclusion criteria: (1) the patients used drugs to inhibit the secretion of gastric acid within 30 days before admission; (2) Specimens used for SARS-CoV-2 nucleic acid testing were collected at an interval of more than 48 hours; (3) cases with incomplete research data; (4) the patients with COVID-19 died in the course of hospitalization.

### Case grouping and requirements

PPIs group: the patients were exposed to PPIs and met the above inclusion and exclusion criteria. A total of 35 patients met the criteria and were included in the PPIs group.

Control group: the patients were not exposed to drugs that inhibited the secretion of gastric acid and met the above inclusion and exclusion criteria. A total of 119 cases met the criteria and were included in the control group.

### Data sources and indicators

The demographic and clinical data was collected from electronic medical record (EMR) including hospital information system (HIS), laboratory information system (LIS) and radiology information system (RIS) of Shanghai Public Health Clinical Center. The demographic and clinical data included age and sex of the patients, the number of pulmonary lobe inflammation by COVID-19 on the CT scan, the onset times, the duration of SARS-CoV-2 clearance after hospitalization and the duration of hospital stay; history of hypertension(HPT), diabetes mellitus(DM), cardiovascular disease(CVD), chronic obstructive pulmonary disease(COPD), chronic liver disease(CLD), chronic gastric disease(CGD), chronic kidney disease(CDK), malignant tumor(MT); laboratory testing results including alanine aminotransferase (ALT), aspartate aminotransferase (AST), creatine kinase (CK), lactate dehydrogenase (LDH), albumin (ALB), prealbumin (PA), creatinine (Cr), D-dimer (D-D), leukocyte (WBC), neutrophils (Neu), lymphocyte(Lym), blood platelet (PLT), and CD4+cells(CD4). All the baseline information was collected in the 24 hours after admission.

The nucleic acid test specimens of SARS-CoV-2 in this study were nasopharyngeal swabs, which were tested by Center of Diseases Prevention and Control of Shanghai before admission, and sampled and tested in the Shanghai Public Health Clinical Center after admission. All the cases admitted to the Shanghai Public Health Clinical Center tested positive for SARS-CoV-2 nucleic acid in Center of Diseases Prevention and Control of Shanghai before hospitalization.

### Statistical analyses

Date are described as median (quartile spacing) or numbers(%). Person chi-square test and fisher exact probability method were used for counting data. The normality of continuous variables was tested by K-S test. T test was used for the differences between data groups conforming to normal distribution, while Manny-Whitney U test was used for the differences between data groups not conforming to normal distribution.

The cumulative probability of SARS-CoV-2 clearance or discharge from COVID-19 cases were conducted through Kaplan-Meier statistics, and the difference was examined by Log-rank test. The possible risk factors of SARS-CoV-2 clearance or discharge from COVID-19 cases were investigated with Cox proportional hazards (PH) regression models for univariate and multivariate analyses to estimate hazard ratios (HRs) and 95% confidence intervals (CIs). PH assumption was verified using Schoenfeld residuals.

In order to reduce the selectivity bias and control the influence of confounding factors, the propensity score method was adopted by logistic regression based on the demographic and clinical indicators in this study. The difference between the two groups was balanced by 1:1 propensity matching.

SPSS software version 23.0 (SPSS Inc. Chicago, IL, USA,) was used for statistical analysis of the data. The P value of two-sided less than 0.05 was considered as statistically significant.

## Results

### Baseline status of COVID-19 cases

A total of 154 COVID-19 cases were included in this study, including 80 males (51.9%), 35 patients (22.7%) in the PPIs group, and 119 patients (77.3%) in the control group. Including: age, sex, hypertension, diabetes mellitus, cardiovascular disease, chronic obstructive pulmonary disease, chronic liver disease, chronic gastric disease, chronic kidney disease, malignant tumor, the number of pulmonary lobe inflammation on the CT scan, the onset times, the duration of SARS-CoV-2 clearance, hospital stays, aminotransferase, aspartate aminotransferase, creatine kinase, lactate dehydrogenase, albumin, prealbumin, creatinine, D - dimer, leukocyte, neutrophils, lymphocytes, platelet and CD4 cells in the two groups had no significant difference; all P > 0.05. See table 1 for details.

**Table 1.** Baseline data among the enrolled cases of COVID-19.

|  | **PPIs group** | **Control group** | **P** |
| --- | --- | --- | --- |
| Cases | 35 | 119 |  |
| Age | 50(35-65) | 52(36-64) | 0.947 |
| Sex,male(%) | 18(51.4) | 62(52.1) | 0.944 |
| HPT(%) | 7(20) | 28(23.5) | 0.661 |
| DM(%) | 2(5.7) | 10(8.4) | 0.870 |
| CVR(%) | 2(5.7) | 5(4.2) | 1.000 |
| COPD(%) | 1(2.9) | 2(1.7) | 0.541 |
| CLD(%) | 1(2.9) | 3(2.5) | 1.000 |
| CGD(%) | 0(0) | 2(1.7) | 1.000 |
| CKD(%) | 2(5.7) | 0(0) | 0.051 |
| MT(%) | 0(0) | 3(2.5) | 1.000 |
| NPLI | 3(2-5) | 4(2-5) | 0.660 |
| The onset times(days) | 3(2-6) | 4(2-7) | 0.418 |
| SARS-CoV-2 clearance(days) | 7(6-9) | 7(6-11) | 0.691 |
| Hospital stays(days) | 21(16-25) | 20(15-26) | 0.507 |
| ALT(U/L) | 21(15-30) | 22(16-35) | 0.643 |
| AST(U/L) | 26(20-42) | 25(20-33) | 0.533 |
| CK(U/L) | 102(55-194) | 80(55-127) | 0.190 |
| LDH(U/L) | 231(193-305) | 219(190-293) | 0.745 |
| ALB(g/L) | 41.0(37.9-44.0) | 41.2(38.1-44.1) | 0.528 |
| PA(mg/L) | 151(93-193) | 158(104-207) | 0.355 |
| Cr(umol/L) | 70.5(52.4-77.3) | 61.3(50.5-77.1) | 0.328 |
| D-D(mg/L) | 0.36(0.28-0.50) | 0.37(0.26-0.64) | 0.828 |
| WBC(*10<sup>9</sup>/L) | 4.4(3.7-5.7) | 4.9(4.0-5.9) | 0.227 |
| Neu(%) | 65.4(55.2-70.6) | 66.6(58.2-72.6) | 0.255 |
| Lym(*10<sup>9</sup>/L) | 1.04(0.73-1.38) | 1.13(0.79-1.52) | 0.196 |
| PLT(*10<sup>9</sup>/L) | 163(121-201) | 176(143-218) | 0.218 |
| CD4(cell/ul) | 411(264-717) | 456(302-650) | 0.711 |

PPIs: proton pump inhibitors; HPT: hypertension; DM-diabetes mellitus; CVD: cardiovascular disease; COPD: chronic obstructive pulmonary disease; CLD: chronic liver disease; CGD: chronic gastric disease; CKD: chronic kidney disease; MT: malignant tumor; NPLI: number of pulmonary lobe inflammation

### Comparison of virus clearance and hospital stay in the baseline status

The duration of SARS-CoV-2 clearance in the PPIs group and the control group was 7(6-9) and 7(6-11) days, respectively. There was no significant difference in the cumulative incidence of SARS-CoV-2 clearance between the two groups, P=0.123. The duration of hospital stay for COVID-19 cases was 21(16-25) days in the PPIs group and 20(15-26) days in the control group. There was no significant difference between the two groups in the cumulative incidence of discharge for COVID-19 cases, P=0.812. See table 1 and figure 2 for details

**FIG 2.**
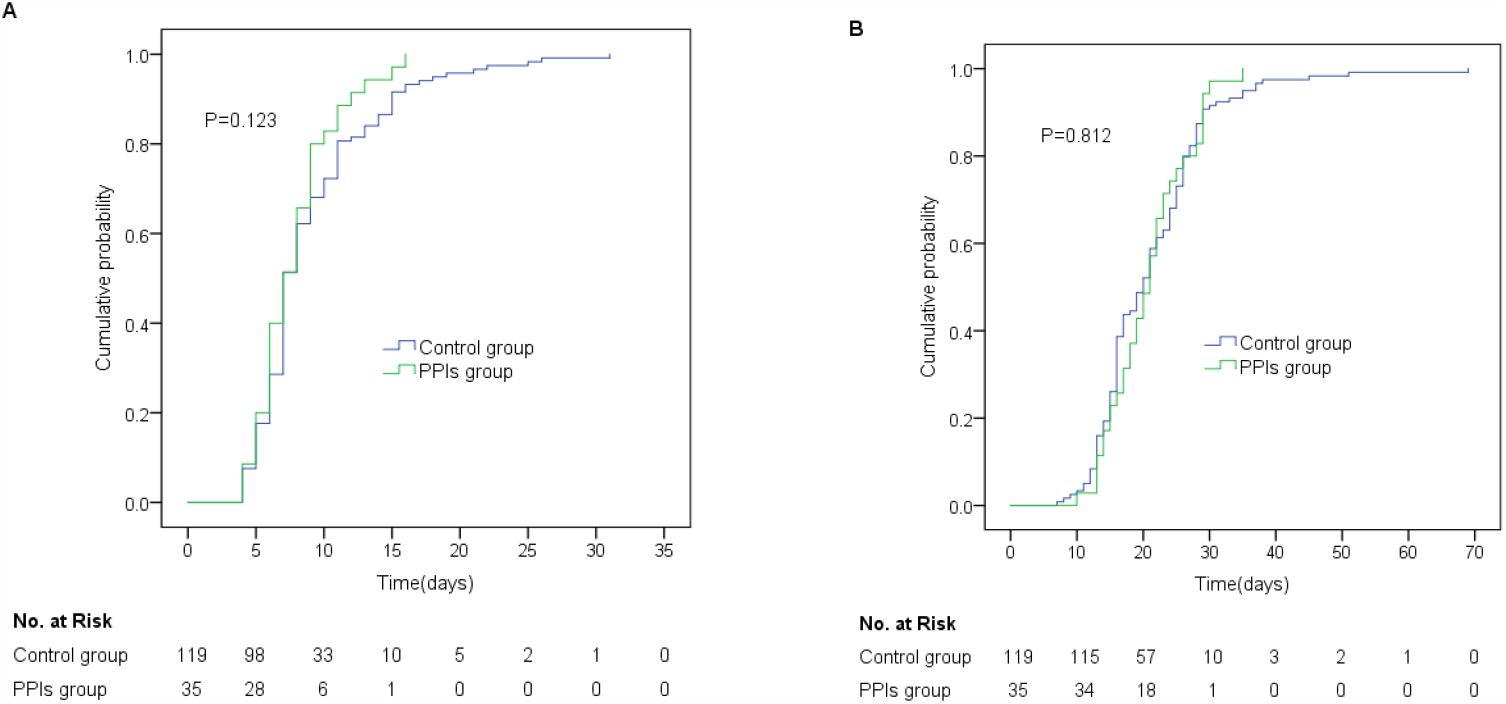
Cumulative probability of SARS-CoV-2 clearance and discharge in COVID-19 patients between PPIs group and control group. Kaplan-Meier curves of (A) SARS-CoV-2 clearance and (B) discharge in the enrolled cases.

### Analysis of the factors influencing viral clearance and hospital stay

#### 1. Analysis of factors affecting SARS-CoV-2 clearance of COVID-19

Univariate analysis: chronic gastropathy prolonged the duration of SARS-CoV-2 clearance in COVID-19 cases, HR 15.202(3.331-69.377), P< 0.001. Multivariate analysis: chronic gastropathy prolonged the duration of SARS-CoV-2 clearance in COVID-19 cases, HR was 20.924(3.547-123.447), P< 0.001. There was no significant difference in the remainder, and all P > 0.05. See table 2 for details.

**Table 2.** Univariate and Multivariate analyses of predictive factors for SARS-CoV-2 clearance in COVID-19 patients.

| Variable | Univariate analysis |  | Mutivariate analysis |  |
| --- | --- | --- | --- | --- |
|  | HR | P | HR | P |
| Age: $\geq 60$/$<60$(years) | 0.837(0.599-1.169) | 0.297 | 0.984(0.645-1.502) | 0.941 |
| Sex: male/female | 0.946(0.686-1.304) | 0.734 | 0.824(0.566-1.200) | 0.313 |
| PPIs/not-PPIs | 1.309(0.893-1.918) | 0.168 | 1.575(0.993-2.499) | 0.054 |
| HPT/not-HPT | 1.187(0.813-1.733) | 0.374 | 1.576(0.959-2.589) | 0.073 |
| DM/not-DM | 0.909(0.503-1.642) | 0.751 | 1.136(0.513-2.517) | 0.754 |
| CVD/not-CVD | 0.596(0.292-1.218) | 0.156 | 0.414(0.161-1.065) | 0.067 |
| COPD/not-COPD | 1.397(0.443-4.403) | 0.568 | 1.344(0.385-4.696) | 0.643 |
| CLD/not-CLD | 2.037(0.750-5.532) | 0.163 | 2.293(0.642-8.193) | 0.201 |
| CGD/not-CGD | 15.202(3.331-69.377) | $< 0.001$ | 20.924(3.547-123.447) | $< 0.001$ |
| CKD/not-CKD | 0.944(0.233-3.821) | 0.935 | 1.105(0.151-8.087) | 0.922 |
| MT/not-MT | 1.144(0.364-3.599) | 0.818 | 0.666(0.155-2.859) | 0.584 |
| NPLI | 1.012(0.913-1.121) | 0.821 | 0.999(0.870-1.146) | 0.985 |
| The onset times(days) | 0.993(0.952-1.035) | 0.735 | 0.993(0.948-1.040) | 0.764 |
| CK: $\geq 198$/$<198$(U/L) | 1.028(0.658-1.606) | 0.904 | 0.908(0.527-1.567) | 0.730 |
| LDH: $\geq 252$/$<152$(U/L) | 1.090(0.785-1.514) | 0.607 | 1.026(0.643-1.635) | 0.916 |
| ALB: $<35$/$\geq 35$(g/L) | 1.259(0.662-2.395) | 0.483 | 0.985(0.408-2.377) | 0.973 |
| PA: $<280$/$\geq 280$(mg/L) | 2.075(0.512-8.410) | 0.307 | 2.812(0.427-18.515) | 0.282 |
| Cr: $\geq 104$/$<104$(umol/L) | 0.933(0.345-2.527) | 0.892 | 0.929(0.270-3.200) | 0.907 |
| D-D: $\geq 0.5$/$<0.5$(mg/L) | 1.053(0.743-1.493) | 0.771 | 1.006(0.601-1.685) | 0.981 |
| WBC |  |  |  |  |
| WBC: $<4$/$4-10$($\times 10^9$/L) | 1.425(0.45-4.517) | 0.547 | 1.414(0.332-6.015) | 0.639 |
| WBC: $>10$/$4-10$($\times 10^9$/L) | 1.251(0.385-4.063) | 0.710 | 1.322(0.31-5.643) | 0.706 |
| Neu: $>70$/$\leq 70$(%) | 1.067(0.754-1.510) | 0.713 | 1.236(0.778-1.964) | 0.370 |
| LYM: $<1.1$/$\geq 1.1$($\times 10^9$/L) | 0.911(0.66-1.257) | 0.570 | 0.91(0.589-1.408) | 0.673 |
| PLT |  |  |  |  |
| PLT: $< 100$/$100-300$($\times 10^9$/L) | 0.713(0.176-2.892) | 0.636 | 1.502(0.277-8.152) | 0.638 |
| PLT: $>300$/$100-300$($\times 10^9$/L) | 0.817(0.164-4.069) | 0.805 | 1.662(0.217-12.737) | 0.625 |
| CD4: $<500$/$\geq 500$(cell/ul) | 0.955(0.689-1.325) | 0.784 | 0.796(0.509-1.246) | 0.319 |
| ALT: $>60$/$\leq 60$(U/L) | 1.709(0.919-3.177) | 0.090 | 1.739(0.865-3.493) | 0.120 |
| AST: $>60$/$\leq 60$(U/L) | 1.717(0.834-3.533) | 0.142 | 1.387(0.466-4.128) | 0.557 |

PPIs: proton pump inhibitors; HPT: hypertension; DM-diabetes mellitus; CVD: cardiovascular disease; COPD: chronic obstructive pulmonary disease; CLD: chronic liver disease; CGD: chronic gastric disease; CKD: chronic kidney disease; MT: malignant tumor; NPLI: number of pulmonary lobe inflammation

#### 2. Analysis of factors affecting the duration of hospital stay of COVID-19

Univariate analysis: chronic liver disease and chronic gastric disease all prolonged the duration of hospital stay in COVID-19 cases, HR were 6.183(2.18-17.534) and 4.155(1.011-17.078), respectively, all P < 0.05. The SARS-CoV-2 clearance shortened the duration of hospital stay in COVID-19 patients, HR was 0.911(0.876-0.948), all P < 0.05. Multivariate analysis: hypertension, chronic obstructive pulmonary disease, chronic liver disease, and malignant tumor prolonged the duration of hospital stay in COVID-19 cases, HR were 1.820 (1.073-3.085), 4.370 (1.205-15.844), 9.011 (2.681-30.290), 5.27 (1.237-22.456), all P < 0.05, respectively. The SARS-CoV-2 clearance shortened the duration of hospital stay in COVID-19 patients, HR was 0.907 (0.869-0.947), all P < 0.05. There was no significant difference in the remainder, and all P > 0.05. See table 2 for details.

### PS-matched analysis of virus clearance and hospital stay

A total of 29 pairs of cases were collected by 1:1 propensity matching, and the corresponding data are shown in table 3. The duration of SARS-CoV-2 clearance in the PPIs group and the control group was 8(6-9) and 8(7-11) days, respectively. There was no significant difference in the cumulative incidence of SARS-CoV-2 clearance between the two groups, P=0.355. The duration of hospital stay for COVID-19 cases was 21(16-27) days in the PPIs group and 19(16-24) days in the control group. There was no significant difference between the two groups in the cumulative incidence of discharge for COVID-19 cases, P=0.817. See table 4 and figure 3 for details.

**Table 3.**
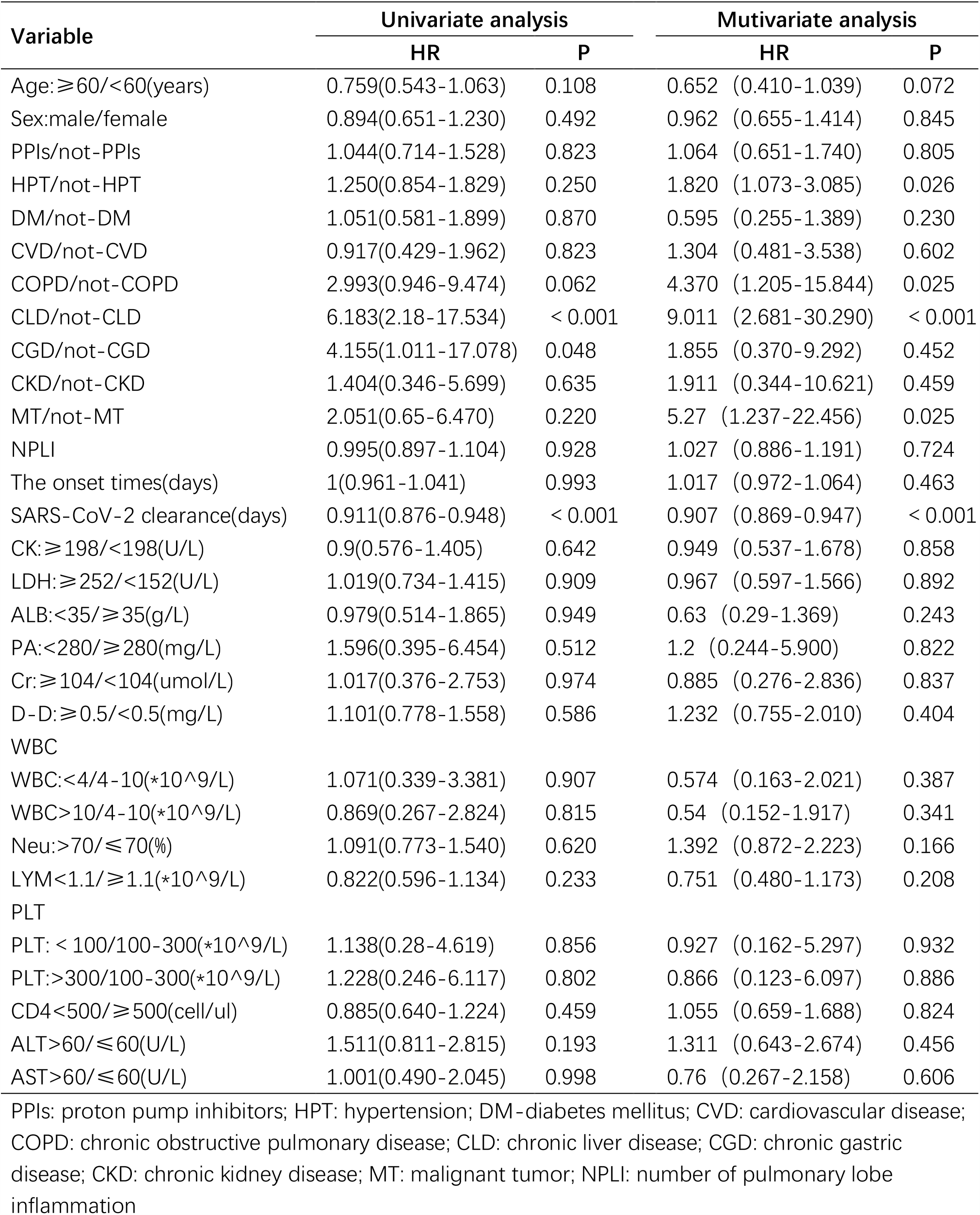
Univariate and Multivariate analyses of predictive factors for discharge in COVID-19 patients.

**Table 4.** Baseline data of the enrolled cases after 1:1 PS-matching analysis.

|  | **PPIs group** | **Control group** | **P** |
| --- | --- | --- | --- |
| Cases | 29 | 29 |  |
| Age | 48(35-67) | 51(36-66) | 0.508 |
| Sex,male(%) | 16(55.2) | 16(55.2) | 1.000 |
| HPT(%) | 4(13.8) | 4(13.8) | 1.000 |
| DM(%) | 1(3.4) | 1(3.4) | 1.000 |
| CVR(%) | 2(6.9) | 2(6.9) | 1.000 |
| COPD(%) | 0(0) | 0(0) | 1.000 |
| CLD(%) | 1(3.4) | 1(3.4) | 1.000 |
| CGD(%) | 0(0) | 0(0) | 1.000 |
| CKD(%) | 0(0) | 0(0) | 1.000 |
| MT(%) | 0(0) | 0(0) | 1.000 |
| NPLI | 4(2-5) | 4(2-5) | 0.923 |
| The onset times(days) | 3(3-7) | 5(2-6.5) | 0.981 |
| SARS-CoV-2 clearance(days) | 8(6-9) | 8(7-11) | 0.616 |
| Hospital stays(days) | 21(16-27) | 19(16-24) | 0.478 |
| ALT(U/L) | 20(15-35) | 20(15-34) | 0.957 |
| AST(U/L) | 27(19-47) | 26(19-46) | 0.852 |
| CK(U/L) | 102(51-181) | 109(51-189) | 0.785 |
| LDH(U/L) | 244(186-326) | 238(185-324) | 0.715 |
| ALB(g/L) | 42.3(37.0-44.2) | 42.6(36.8-44.4) | 0.969 |
| PA(mg/L) | 153(92-192) | 155(89-201) | 0.750 |
| Cr(umol/L) | 64.9(51.5-80.6) | 64.3(51.2-80.2) | 0.797 |
| D-D(mg/L) | 0.34(0.27-0.61) | 0.33(0.27-0.52) | 0.744 |
| WBC(*10<sup>9</sup>/L) | 4.3(3.8-6.0) | 4.4(3.9-5.9) | 0.864 |
| Neu(%) | 62.8(52.5-73.3) | 63.1(52.15-73.0) | 0.864 |
| Lym(*10<sup>9</sup>/L) | 1.07(0.70-1.40) | 1.04(0.7-1.39) | 0.901 |
| PLT(*10<sup>9</sup>/L) | 165(119-202) | 167(119-204) | 0.981 |
| CD4(cell/ul) | 406(236-732) | 403(229-724) | 0.932 |

PPIs: proton pump inhibitors; HPT: hypertension; DM-diabetes mellitus; CVD: cardiovascular disease; COPD: chronic obstructive pulmonary disease; CLD: chronic liver disease; CGD: chronic gastric disease; CKD: chronic kidney disease; MT: malignant tumor; NPLI: number of pulmonary lobe inflammation

**FIG 3.**
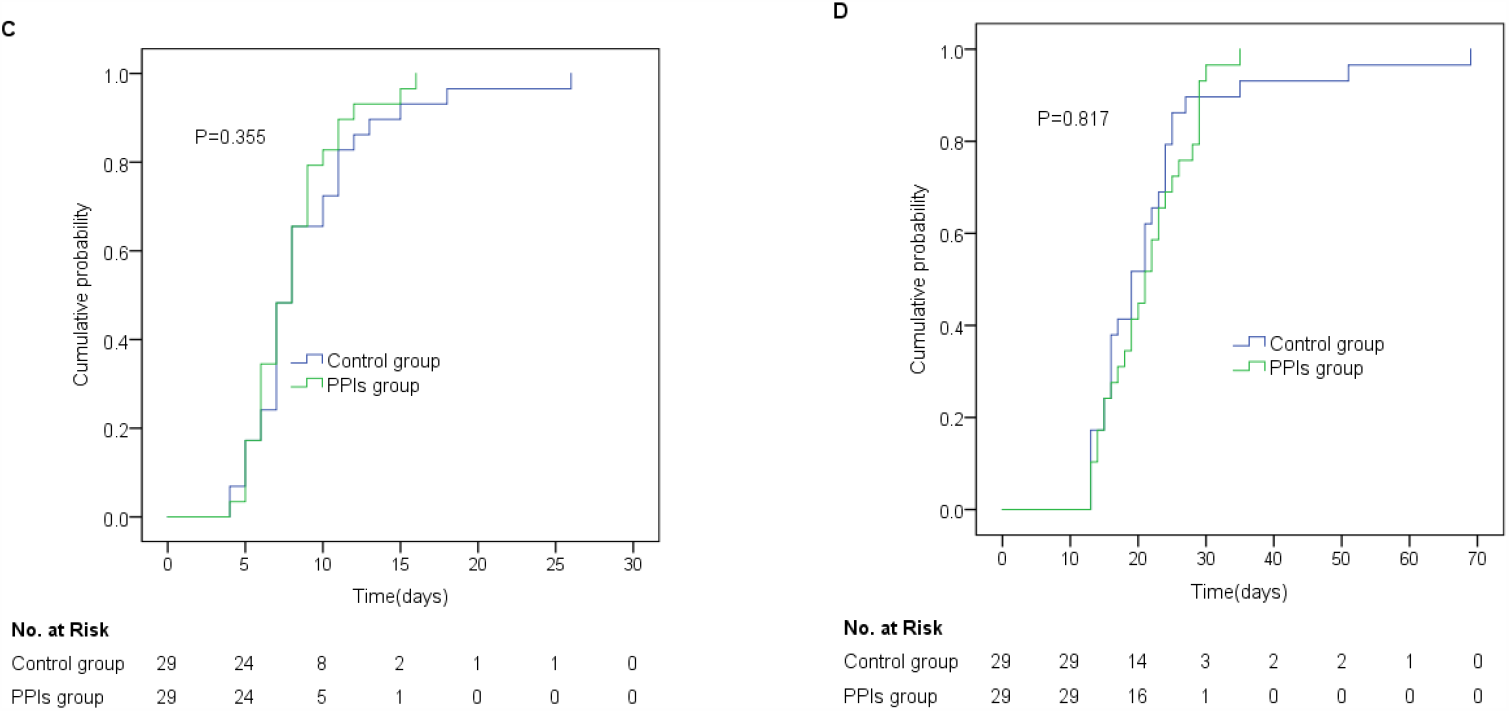
Cumulative probability of SARS-CoV-2 clearance and discharge in COVID-19 patients between PPIs group and control group by 1:1 PS-matching analysis. Kaplan-Meier curves of (C) SARS-CoV-2 clearance and (D) discharge by 1:1 PS-matching analysis.

## Discussion

Patients with gastric acid-related diseases often need to take PPIs for treatment. However, the risk of using PPIs on the occurrence of pneumonia have been reported inconsistently, and the effect of PPIs on the course of pneumonia has not been reported. Especially in this COVID-19 outbreak, it is difficult to draw on appropriate references regarding the safety or risk of PPIs in COVID-19 cases. Currently, most of the existing studies on the safety or risk of PPIs use are retrospective cohort studies or case-control studies. The mechanism of the safety or risk of PPIs has not been fully clarified, and more high-quality design studies are needed to confirm it. This study can not only preliminarily determine the effect of PPIs on the course of COVID-19, but also further provide the basis to verify whether the PPIs use increases the incidence of pneumonia.

To date, no effective drug treatment for COVID-19 has been found in large sample of high-quality clinical studies[20], except for remdesivir superior to placebo in shortening recovery times in adult patients hospitalized with COVID-19 and evidence of lower respiratory tract infection[21]. Therefore, other drugs were not included in the statistical analysis.

### Key findings

In this study, we found that PPIs use in common COVID-19 did not prolong or shorten the duration of SARS-CoV-2 clearance or hospital stay. After preliminary screening, chronic gastropathy prolonged SARS-CoV-2 clearance, while hypertension, chronic bronchitis, chronic liver disease, and malignant tumors might prolong the duration of hospital stay in COVID-19 patients, the SARS-CoV-2 clearance would shorten the duration of hospital stay in COVID-19 patients. Primary diseases and comorbidity are the main factors affecting the course of COVID-19, and the etiological treatment is important in COVID-19.

### Comparison with related literature

So far, there have been no reported effects of PPIs on COVID-19. However, there have been more studies on the effect of PPIs on the occurrence of community acquired pneumonia or hospital acquired pneumonia, but these reports are inconsistent.

several studies suggested that PPIs increased the incidence of community acquired pneumonia,[5-9] and it also showed a statistically significant risk of the incidence of hospital acquired pneumonia.[8-9]Studies reported that the occurrence of pneumonia is related to the duration of proton pump inhibitor use, and its short-term use increased the risk of pneumonia.[5,10-11] The risk of pneumonia was related to the dose of proton pump inhibitors, which were found to have a higher risk of pneumonia at high doses. [5,11-12] On the other side, the studies suggested that PPIs did not increase the incidence of pneumonia. They suggested that the increased incidence of pneumonia caused by PPIs was due to confounders, and proton pump itself was not an independent factor affecting the occurrence of pneumonia.[13-16] According to our study, the PPIs use has no relation with the duration of SARS-CoV-2 clearance and hospital stay by Cox regression and the propensity score method. We prefer the opinion that the PPIs use does not increase the probability of pneumonia. the PPIs use were used at short and conventional doses in our study cases, so we cannot provide evidence to assess whether high doses or long-term use of PPIs increase the risk of pneumonia.

Basic studies have shown that PPIs can effectively inhibit virus-specific serine proteases and thus play an antiviral role.[17] And H+/K+-ATPase inhibitors can be tried to treat viral infection and respiratory disease.[18] However, other findings have been reported that gastrointestinal microorganisms modulate immune responses at distant mucosal sites and have the ability to significantly influence mortality during respiratory viral infection.[19] This is inconsistent with our research on the effects of PPIs use on COVID-19. In our study, the PPIs use did not shorten the duration of SARS-CoV-2 clearance, and there was no death in the proton pump inhibitor group. This may be related to the virus species, and more basic studies and clinical studies needed to confirm the effects of proton pump inhibitors use on various virus.

### strengths of study

The data information included in this study was very complete, the study date was continuously monitored, including demographic characteristics and clinical information. Cox regression model was used to determine whether PPIs use affected the course of COVID-19, effectively excluding the influence of confounders. The effect of PPIs use on the course of COVID-19 was further validated by propensity matching and long-rank test. Therefore, the results are relatively reliable. The study was conducted on COVID-19, which effectively excluded the confounding effects of pneumonia caused by other pathogens. It also makes up for the lack of pathogen parameters in the baseline data from previous studies, which was the relation between PPIs use and pneumonia occurrence.

### limitations of study

The baseline data of our study lacked the viral load of SARS-CoV-2, but lymphocytes and CD4+ cells were included as the baseline reference and matching analysis was performed to make up for the deficiencies in our study, because it was proved that SARS-CoV-2 would affect the change of lymphocyte and CD4+ cell counts[22-24]. This study included only common COVID-19, due to the low number of severe and critical COVID-19 cases, and severe and critical cases were not included in the study; therefore, the study does not fully represent the effect of proton pump inhibitors on the course of severe or critical COVID-19. Since the subjects we included in this study are common COVID-19 cases, and PPIs have been used for a short time and at a conventional dose, so the study cannot represent the long-term or high-dose impact of PPIs on the course of COVID-19.

## Conclusions

The use of proton pump inhibitors does not prolong or shorten the duration of SARS-CoV-2 clearance or hospital stay. Therefore, the appropriate use of proton pump inhibitors in the treatment or prevention of the related diseases will not affect the course of COVID-19.

## Data Availability

This study was supported by the Shanghai Public Health Clinical Center for data access.

## APPENDIX

### Author Contributions

Conceptualization, Xiao-Yu Zhang, Hai-Bing Wu, Yun Ling, Zhi-Ping Qian and Liang Chen; Formal analysis, Xiao-Yu Zhang; Supervision, Liang Chen; Writing – original draft, Xiao-Yu Zhang; Writing – review & editing, Xiao-Yu Zhang, Hai-Bing Wu, Yun Ling, Zhi-Ping Qian and Liang Chen. All authors have read and agreed to the published version of the manuscript.

### Funding

This research received no external funding.

## Acknowledgments

This study was supported by the Shanghai Public Health Clinical Center for data access.

## Ethical approval

Informed consents of patients were obtained for diagnosis and treatment, and the study. Protocol was approved by the Shanghai Public Health Clinical Center Clinical Committee. All the data received Institutional Review Board (IRB) approval by the Ethics Committee. The IRB number was YJ-2020-S015-01.

